# Cross-EHR validation of antidepressant response algorithm and links with genetics of psychiatric traits

**DOI:** 10.1101/2024.09.11.24313478

**Authors:** Julia M. Sealock, Justin D. Tubbs, Allison M. Lake, Peter Straub, Jordan W. Smoller, Lea K. Davis

**Author notes:** <u>Corresponding Author Information:</u> Lea K. Davis, PhD, Division of Data-Driven and Digital Medicine, Department of Medicine Charles Bronfman Institute for Personalized Medicine, The Hamilton and Amabel James Center for Artificial Intelligence, 3 E. 101st St., New York, NY 10029.

## Abstract

**Objective:** Antidepressants are commonly prescribed medications in the United States, however, factors underlying response are poorly understood. Electronic health records (EHRs) provide a cost-effective way to create and test response algorithms on large, longitudinal cohorts. We describe a new antidepressant response algorithm, validation in two independent EHR databases, and genetic associations with antidepressant response.

**Method:** We deployed the algorithm in EHRs at Vanderbilt University Medical Center (VUMC), the All of Us Research Program, and the Mass General Brigham Healthcare System (MGB) and validated response outcomes with patient health questionnaire (PHQ) scores. In a meta-analysis across all sites, worse antidepressant response associated with higher PHQ-8 scores (beta = 0.20, p-value = 1.09 x 10^−18^).

**Results:** We used polygenic scores to investigate the relationship between genetic liability of psychiatric disorders and response to first antidepressant trial across VUMC and MGB. After controlling for depression diagnosis, higher polygenic scores for depression, schizophrenia, bipolar, and cross-disorders associated with poorer response to the first antidepressant trial (depression: p-value = 2.84 x 10^−8^, OR = 1.07; schizophrenia: p-value = 5.93 x 10^−4^, OR = 1.05; bipolar: p-value = 1.99 x 10^−3^, OR = 1.04; cross-disorders: p-value = 1.03 x 10^−3^, OR = 1.05).

**Conclusions:** Overall, we demonstrate our antidepressant response algorithm can be deployed across multiple EHR systems to increase sample size of genetic and epidemiologic studies of antidepressant response.

## Introduction

Antidepressants are first-line medications to treat psychiatric disorders including depression, anxiety, and obsessive-compulsive disorder, and are among the most widely prescribed medications in the United States(1–3). However, only 35% of depression patients will reach remission with their first antidepressant trial, and around 22% of depression patients will fail to respond to multiple antidepressant trials(4–7). There are currently few tools to assist clinicians in identifying the most effective treatment for a patient, resulting in trial-and-error for antidepressant prescribing.

Studies focused on identifying factors associated with antidepressant response found inconsistent associations with inflammatory biomarkers, brain anatomy and function, sex, and adverse life events(8–13). Pharmacogenetic studies and clinical guidelines for antidepressants focus on drug metabolism, particularly of SSRIs, with few replicated associations between candidate genes and response(14,15). Genome-wide association studies (GWAS) of antidepressant response reveal evidence for a complex genetic architecture of response, remission, and treatment resistance(16–19). However, most studies suffer from low sample sizes, hindering power to detect associations and generalize results. Better understanding of factors influencing antidepressant response, including symptomology, biomarkers, and genetics, could help improve treatment outcomes and guide the development of new treatments.

A major limitation for identifying factors affecting treatment response is a scalable and longitudinal response algorithm. Traditional epidemiologic studies of antidepressant response typically have low sample sizes, minimal follow-up time, and examine a limited number of antidepressants(4,9,12,20). More recently, several algorithms using machine learning techniques were applied to electronic health records (EHRs), which contain longitudinal information on patient care, including diagnoses, medications, and demographic information(21,22). Although ML models applied to EHR data achieved high levels of accuracy (AUC: 70 – 76%), portability across healthcare systems is limited due to restricted access to clinical notes in research EHRs, sparse documentation of response, and the computational burden of implementing ML in a new EHR dataset(23,24).

In this study, we utilized medication information in EHRs to create a new rule-based antidepressant response algorithm based on medication additions and switching. Rather than relying on clinical notes, we used structured data to infer response for all individuals with documented use of antidepressants. We validated our algorithm using depression severity scores from the Patient Health Questionnaire (PHQ) across three EHR systems. Using genetic approaches, we identify a genetic component to antidepressant response and describe correlations between response and polygenic risk of psychiatric traits.

## Methods

Descriptions of Vanderbilt University Medical Center (VUMC), Mass General Brigham Biobank (MGB) and the All of Us Research Program can be found in the Supplemental Methods. Data collection was approved by Institutional Review Boards for each site, and all participants provided written informed consent for inclusion in the respective institutional biobanks for broad-based research.

### Antidepressant response algorithm

We extracted antidepressants (including SSRIs, SNRIs, TCAs, atypical antidepressants, and MAOIs) and second-generation antipsychotics from EHRs at VUMC and manually mapped medications to generic names after removing dosage and route information from medication names (Supplementary Tables 1 & 2). Medications were extracted from various sources, including medications dispensed from the VUMC pharmacy, in-patient administered medications, patient reported medications, procedural administration, written prescriptions, medications listed in clinical notes, and physician administered drugs.

Our algorithm was adapted from Fabbri et al’s algorithm (ref. 18) to identify treatment resistant depression cases from EHRs from the UK Biobank and the Extended Cohort for E-health, Environment and DNA (EXCEED). Rather than identifying treatment resistant depression, our algorithm outputs response to each antidepressant extracted from an EHR record. To ensure antidepressants were prescribed to treat the same depressive episode we grouped antidepressants by prescription episodes. Because antidepressants are frequently prescribed prior to the assignment of a depression diagnosis, we did not require a diagnosis of depression for samples included in our algorithm. Prescription episodes were defined using gaps in antidepressant records as in Fabbri et al 2021. If a gap > or = 24 weeks between antidepressant records, was detected, the time period after the gap was considered a new prescription episode.

For each antidepressant, we required an adequate antidepressant trial defined as evidence of the same medication being prescribed for at least 6 consecutive weeks from the initial prescription, the minimum time given in the United States for antidepressants to show maximal efficacy. Second generation antipsychotics can be used to augment antidepressants in the treatment of major depression(25). To model antipsychotic use, we extracted antipsychotics used during antidepressant treatment and required evidence of antipsychotic use for at least two weeks.

Within each prescription episode, we determined whether a patient switched to a different antidepressant or if an additional antidepressant was added in the same episode without evidence of discontinuation. We determined antidepressant response by combining the information on antidepressant additions and switches with the dates of antipsychotic use. If an individual switched from an antidepressant, we labelled the antidepressant trial as “non-responder”. If an individual had multiple antidepressants but did not have antipsychotics in the episode, we labelled the trial as “intermediate – antidepressants only”. If an antipsychotic was added during a prescription episode, we labelled the trial as “intermediate – with + antipsychotics”. Finally, if an individual did not have evidence of adding antidepressants, switching from an antidepressant, or augmentation with antipsychotics, we labelled the trial as “responder”. As a result, our antidepressant algorithm outputs response classifications of “responder”, “intermediate – antidepressants only”, “intermediate – with + antipsychotics”, and “non-responder” for each antidepressant trial in an individual’s EHR.

**Figure 1.**
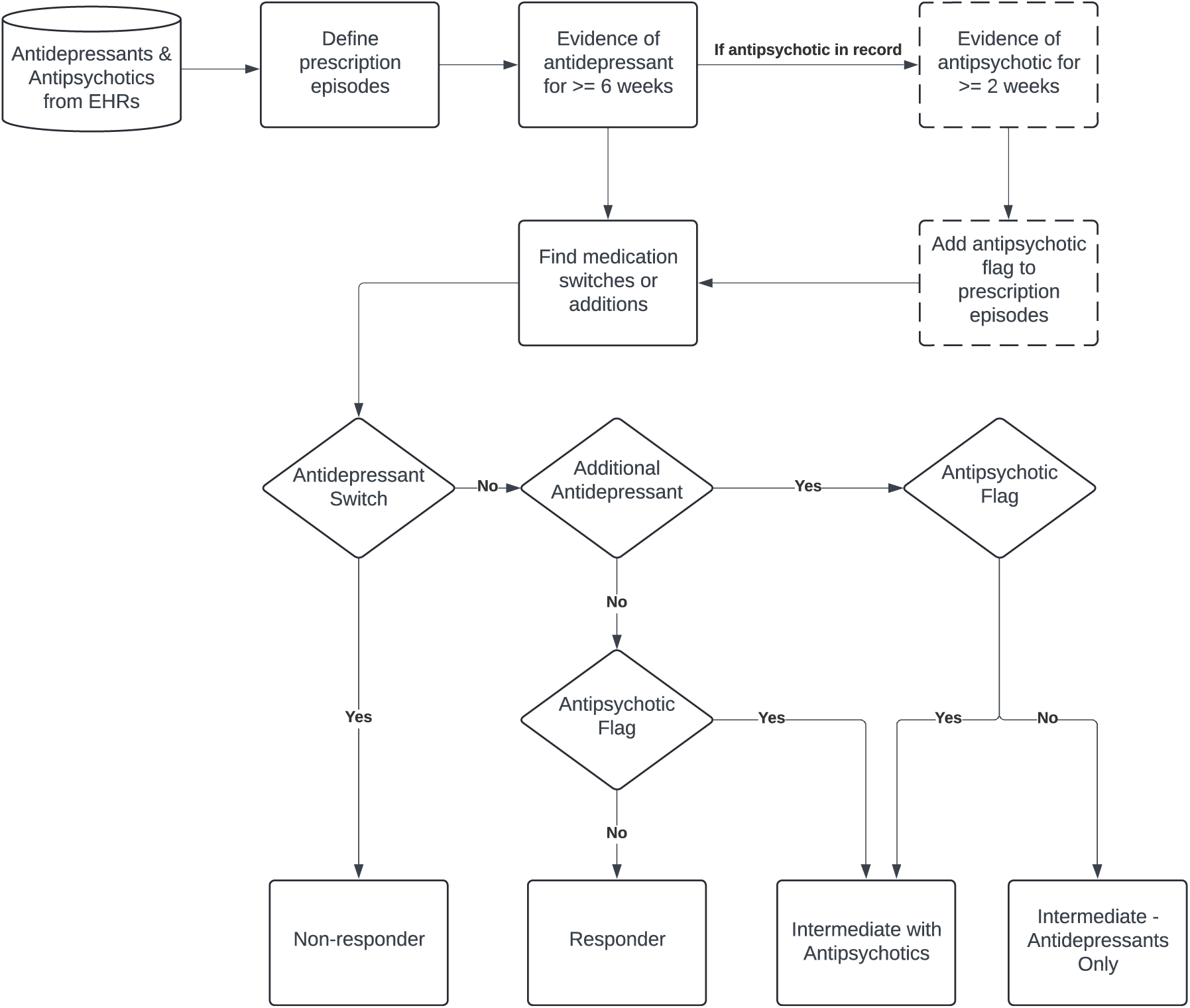
Overview of the antidepressant response algorithm used to infer response from medication switching and additions. Individuals without any medication switches or additions were designated as ‘responder’. Individuals without a switch but with an added antidepressant were labelled ‘intermediate – antidepressants only’. Individuals without a switch but with an added antipsychotic were labelled ‘intermediate with antipsychotics’. individuals with evidence of switching from an antidepressant and starting another were labelled ‘non-responders’.

### Validation using Patient health questionnaire (PHQ)

A subset of VUMC patients (N = 106,644) have one or more recorded responses for the patient health questionnaire (PHQ), a validated screener for depression symptoms and severity(26,27). The PHQ data at VUMC primarily consisted of PHQ-2 (N samples = 72,435, N entries = 120,577) and PHQ-8 (N samples = 45,396, N entries = 122,041), which contain two and eight questions on depression symptoms, respectively. We conducted minimal cleaning on the PHQ values by removing values greater than the maximum possible score for each version (PHQ-2_max_ = 6, PHQ-8_max_ = 24). If an individual had multiple measurements of the same PHQ version on the same date, we used the average score of all measurements. Cleaning resulted in 119,850 PHQ-2 entries on 72,389 individuals, and 121,516 PHQ-8 entries on 45,394 individuals.

We validated our antidepressant response algorithm by testing whether changes in PHQ associated with the rule-based response classifications. To find corresponding response variables for each PHQ entry, we first filtered response outcomes to the closest date (within a maximum window of two weeks) for each PHQ entry. If a PHQ measurement did not have an antidepressant response outcome within two weeks of the PHQ assessment, it was removed from analysis. We matched PHQ-8 and PHQ-2 measurements to antidepressant responses independently from one another, creating two datasets, a PHQ-8 response-matched and a PHQ-2 response-matched dataset. Within each PHQ dataset, we required individuals to have more than one PHQ-response-matched entry for inclusion, leaving 4,916 PHQ-8 assessments collected on 1,493 patients and 5,936 PHQ-2 assessments collected on 2,378 patients. For VUMC, the median number of observations in the model was 3 for PHQ-8 with a range of 2 – 14 observations. For PHQ-2 the median number of observations in the model was 2 with a range of 2 – 59 (Supplementary Table 3, Supplementary Figure 1).

PHQ-2 and PHQ-8 scores were z-score scaled within version so that the resulting beta estimates can be interpreted per standard deviation increase in PHQ-2 or PHQ-8 score. We then fitted multivariable linear mixed regression models which tested the association between z-score scaled PHQ values and antidepressant response, including covariates for EHR-reported race and sex, age at PHQ measurement, and a random intercept to control for the baseline PHQ score. For the regression, we transformed the beta estimate to represent the change in PHQ score by multiplying the beta estimate by the standard deviation of the untransformed PHQ scores (Supplementary Table 4). We performed the regression for PHQ-2 and PHQ-8 in all samples and stratified by to depression cases, defined as the presence of 2 or more ICD codes for major depression, depression, adjustment reaction, or dysthymic disorder.

### Replication in the All of Us Research Program and the MGB Healthcare System

To validate our algorithm in two independent EHR datasets, we used data from the All of Us Research Program (data release version 7)(28) and the Mass General Brigham healthcare system (MGB). From the MGB and All of Us datasets, we extracted PHQ-2 and PHQ-9 measurements to replicate the validation framework as described above. In the All of Us cohort, PHQ scores were extracted from EHRs. To maintain analysis of EHR data, PHQ scores recorded in the COVID-19 Participant Experience survey (COPE) were not considered in our analyses. PHQ scores were z-score scaled within site and PHQ version to enable meta-analysis. We conducted meta-analysis of the effect sizes across the three sites using a fixed effects inverse weighted analysis. PHQ-2 scores from all sites were meta-analyzed together. Because of the similarity of questions and scoring range for PHQ-8 and PHQ-9, the PHQ-8 scores VUMC were meta-analyzed with the PHQ-9 scores from All of Us and MGB using a fixed-effects inverse weighted meta-analysis method.

### Genome-wide association scan for first trial antidepressant response

We conducted an ordinal genome-wide association scan (GWAS) of response to first antidepressant trial within individuals of European ancestry who also had genetic information available in BioVU (N = 30,152). We defined first trial response as the last recorded response within the first prescription episode for the first recorded antidepressant. We combined responses from all antidepressant classes to increase power and to find genetic risk factors relevant to the first recorded trial. Most samples were prescribed SSRIs as first documented trial (N = 17,345), followed by SNRI (N = 4,956), TCA (N = 4,243), atypical antidepressants (N = 3,593), and MAOIs (N = 15) (Supplementary Table 5).

Quality control on imputed genotyping data was completed as previously described(29). We used the Julia package, OrdinalGWAS, to conduct a likelihood ratio test on SNP dosages(30) for SNPS with a minor allele frequency >= 0.1%. Response was ordered from lowest to highest as ‘responder’, ‘intermediate – antidepressants only’, ‘intermediate with antipsychotics’, and ‘non-responder’. The GWAS was covaried for sex, age at response and top 10 principal components of ancestry.

We calculated the genetic correlation between antidepressant response and psychiatric traits using High-Definition Likelihood (HDL)(31). HDL is a genetic correlation method that fully accounts for linkage disequilibrium across the genome to reduce variance in correlation estimates and increase power when using GWAS with low sample sizes. We calculated genetic correlation with the largest available summary statistics for depression, schizophrenia, anxiety, bipolar, and cross-disorders(32–36). We estimated the heritability of the GWAS of antidepressant response using HDL.

### Associations with psychiatric polygenic scores

In VUMC and MGB, we calculated polygenic scores (PGS) for anxiety, depression, bipolar, schizophrenia, and cross-disorders for individuals of European and African ancestry groups, separately, using PRS-CS and SNP weights from the largest available meta-analyses(37). PRS-CS-auto uses a Bayesian framework to model linkage disequilibrium (LD) from an external reference panel and applies a continuous shrinkage prior on SNP effect sizes which is automatically learnt from the data. For summary statistics derived from European ancestries, the LD reference panel was constructed from European samples in the 1000 Genomes Project phase 3 (1KGP) and applied to both European and African ancestry samples in BioVU. To calculate schizophrenia PGS in individuals of African ancestry in BioVU, we used weights derived from an African ancestry GWAS(36) and an LD reference panel constructed from African samples in the 1KGP. PGS were scaled to have a mean of zero and a unit standard deviation (SD) so that effect estimates in subsequent analyses are interpreted per 1 SD increase in PGS.

We tested for associations between genetic predisposition to depression, bipolar disorder, and schizophrenia, and response to first antidepressant trial using an ordinal regression. Ordered response outcomes were ranked from lowest to highest – ‘responder’, ‘intermediate – antidepressants only’, ‘intermediate with antipsychotics’, and ‘non-responder’. All ordinal regression models were controlled for sex, age at response, and top 10 principal components of ancestry. Lastly, we conducted a sensitivity analysis including an additional covariate for depression diagnosis, defined as the presence of a phecode for either major depression (296.22), depression (296.2), adjustment reaction (304), or dysthymic disorder (300.4). To define a phecode, we required at least two instances of component ICD codes on different days (Supplemental Table 6). Results from VUMC and MGB were meta-analyzed using a fixed-effects inverse weighted approach for each trait. Analyses with and without a depression covariate were meta-analyzed separately.

## Results

### Description of algorithm results

We applied our antidepressant response algorithm to 671,851 individuals with 20,298,966 antidepressant records in VUMC, resulting in 315,935 individuals with 7,152,656 antidepressant response outcomes (Table 1). The median age of first antidepressant mention was 47.9 years (IQR: 31.9 – 61.2) and the median time between first and last antidepressant record was 10.4 months (IQR: 3.9 – 43.0). Most individuals had an SSRI for their first recorded antidepressant trial (57.4%), followed by SNRIs (16.3%), TCAs (14.0%), Atypical antidepressants (12.3%), and MAOIs (0.03%). Among all antidepressant users, 33,851 (10.7%) had concurrent antipsychotic use, including 26,290 individuals with antipsychotic use recorded in their first antidepressant episode. After restricting to first recorded antidepressant trial, 244,807 (77.5%) individuals were identified as responders, 22,923 (7.3%) were intermediate – antidepressant only, 21,397 (6.7%) were intermediate with antipsychotics, and 26,808 (8.5%) were non-responders.

**Table 1.** Characteristics of response to first EHR-documented antidepressant trial in VUMC, All of Us, and MGB.

| Metric | Cohort | Responder | Intermediate – antidepressants only | Intermediate – with antipsychotics | Non-responder |
| --- | --- | --- | --- | --- | --- |
| Total | VUMC | 244,807 | 22,923 | 21,397 | 26,808 |
|  | All of Us | 39,972 | 3,545 | 3,922 | 6,796 |
|  | MGB | 221,570 | 19,362 | 21,071 | 23,033 |
| N Female (%) | VUMC | 164,342 (67.1%) | 16,373 (71.4%) | 12,239 (57.2%) | 18,339 (68.4%) |
|  | All of Us | 27,879 (69.7%) | 2,464 (69.5%) | 2,283 (58.2%) | 4,607 (67.8%) |
|  | MGB | 150,215 (67.8%) | 13,339 (68.9%) | 11,880 (56.4%) | 15,490 (67.3%) |
| N EHR-reported White (%) | VUMC | 206,963 (84.5%) | 19,848 (86.6%) | 17,740 (82.9%) | 23,062 (86%) |
|  | All of Us | 26,558 (66.4%) | 2,498 (70.5%) | 2,020 (51.5%) | 4,691 (69.0%) |
|  | MGB | 183,485 (82.8%) | 16,397 (84.7%) | 17,130 (81.3%) | 18,530 (80.4%) |
| Median age at first antidepressant, years (IQR) | VUMC | 48.1 (31.9 - 61.6) | 50.3 (37.4 - 61.1) | 46.3 (27.9 - 59.9) | 45.4 (30 - 58.5) |
|  | All of Us | 51.3 (38.5 - 61.2) | 50.3 (38.5 - 59.5) | 48.0 (37.1 - 55.9) | 47.1 (34.9 - 56.8) |
|  | MGB | 50.6 (35.8 – 63.6) | 51.1 (37.9 – 62.2) | 48.0 (30.9 – 61.1) | 45.5 (31.4 – 58.1) |
| N Depression Cases (%) | VUMC | 44,343 (18.1%) | 6,668 (29.1%) | 6,707 (31.3%) | 11,990 (44.7%) |
|  | All of Us | 27,429 (68.6%) | 2,935 (82.8) | 3,323 (84.7) | 5,755 (84.7%) |
|  | MGB | 140,533 (63.4%) | 15,387 (79.5%) | 17,139 (81.3%) | 19,280 (83.7%) |
| N SSRI for first antidepressant (%) | VUMC | 143,824 (58.7%) | 9,425 (41.1%) | 12,473 (58.3%) | 15,487 (57.8%) |
|  | All of Us | 22,259 (55.7%) | 1,626 (45.9) | 2,225 (56.7%) | 3,964 (58.3%) |
|  | MGB | 134,083 (60.5%) | 9,087 (46.9%) | 12,288 (58.3%) | 13,230 (57.4%) |
| N SNRI for first antidepressant (%) | VUMC | 39,544 (16.2%) | 3,783 (16.5%) | 4,213 (19.7%) | 4,041 (15.1%) |
|  | All of Us | 6,399 (16.0%) | 536 (15.1%) | 681 (17.4) | 885 (13.0%) |
|  | MGB | 21,989 (9.9%) | 2,232 (11.5%) | 3,025 (14.4%) | 2,275 (9.9%) |
| N TCA for first antidepressant (%) | VUMC | 34,236 (14.0%) | 5,022 (21.9%) | 1,218 (5.7%) | 3,847 (14.4%) |
|  | All of Us | 5,431 (13.6%) | 467 (13.2%) | 183 (4.7%) | 860 (12.7%) |
|  | MGB | 35,298 (15.9%) | 2,596 (13.4%) | 1,015 (4.8%) | 3,572 (15.5%) |
| N Atypical for first antidepressant (%) | VUMC | 27,138 (11.1%) | 4,690 (20.5%) | 3,467 (16.2%) | 3,419 (12.8%) |
|  | All of Us | 5861 (14.7%) | 915 (25.8%) | 827 (21.1%) | 1,086 (16.0%) |
|  | MGB | 30,097 (13.6%) | 5,442 (28.1%) | 4,697 (22.3%) | 3,939 (17.1%) |
| N MAOI for first antidepressant (%) | VUMC | 65 (0%) | 3 (0%) | 26 (0.1%) | 14 (0.1%) |
|  | All of Us | 22 (0.1%) | < 20 | < 20 | < 20 |
|  | MGB | 103 (0%) | 5 (0%) | 46 (0.2%) | 17 (0.1%) |

### Validation with patient health questionnaire

Changes in PHQ-8 significantly associated with algorithm-defined antidepressant response across all samples (beta = 0.35, SE = 0.05, p-value = 1.28 x 10^−15^), with worse antidepressant response associating with higher PHQ-8 scores (Figure 2). The transformed beta estimate corresponded to a 1.88-point increase in PHQ-8 with each increase in worsening antidepressant response. The association remained after restricting to depression cases and corresponded to a 1.73-point increase in PHQ-8 scores with worsening response (beta = 0.31, SE = 0.08, p-value = 4.30 x 10^−6^) (Figure 2, Table 2).

**Figure 2.**
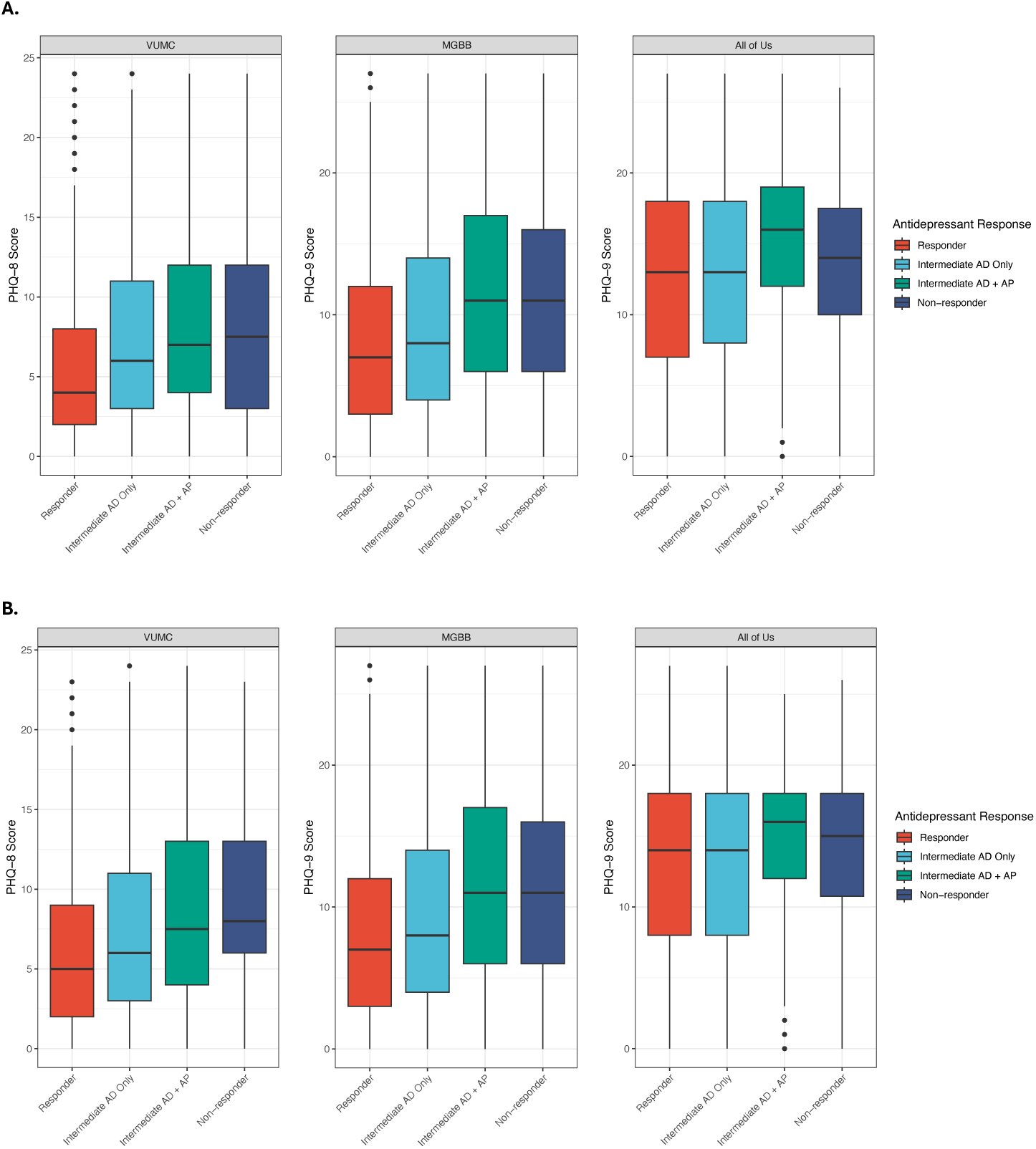
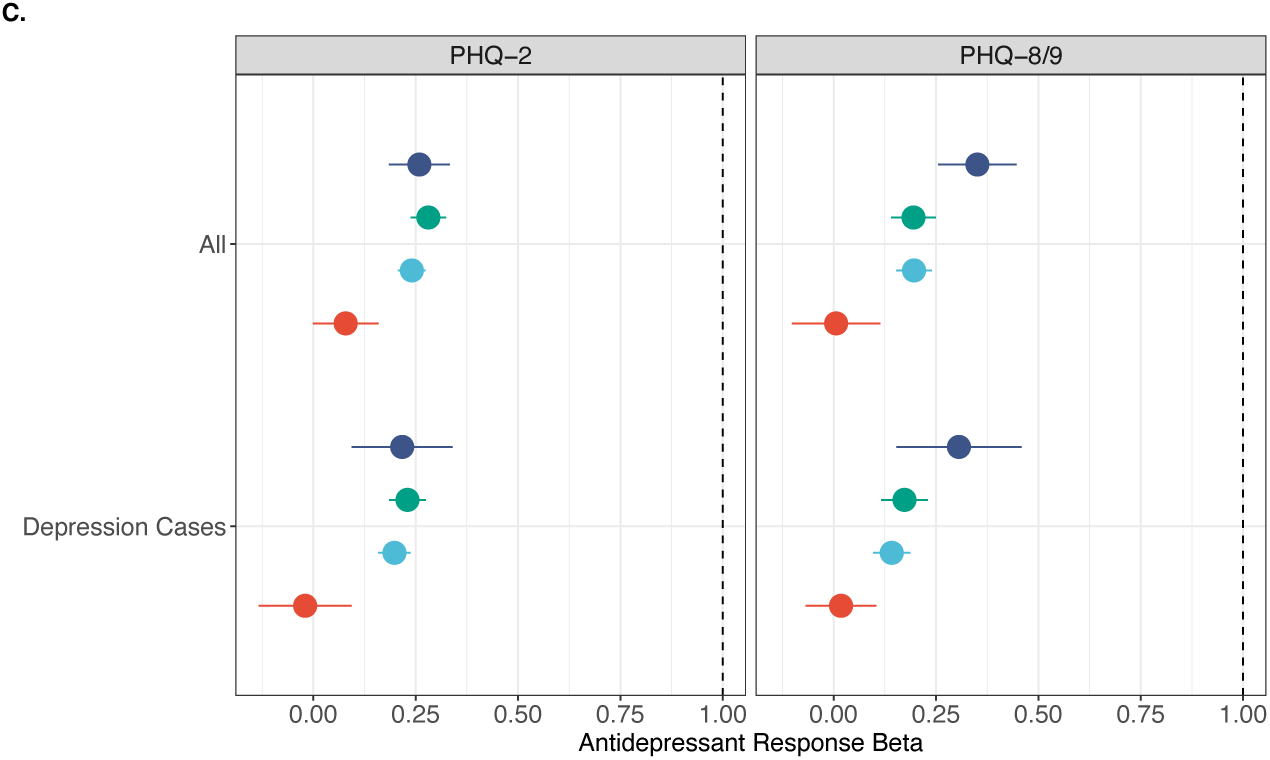
**A**. Patient health questionnaire scores by antidepressant response in all individuals and **B.** stratified to depression cases in VUMC, All of Us, and MGB. VUMC results are with PHQ-8; All of Us and MGB results are with PHQ-9. Intermediate AD only = intermediate – antidepressants only; Intermediate AD + AP = intermediate with antipsychotics. **C**. Cohort-specific and meta-analyzed effect estimates of antidepressant response on PHQ scores from longitudinal models for each PHQ version.

**Table 2.** Results of linear mixed model between patient health questionnaire scores and antidepressant response outcomes in VUMC, All of Us, MGB, and the meta-analysis. Scores from each PHQ version were tested separately. Sensitivity analyses were conducted within depression cases.

| Cohort | Sample | Version | p-value | Beta | SE | N Observations | N Individuals |
| --- | --- | --- | --- | --- | --- | --- | --- |
| VUMC | All | PHQ-8 | 1.28E-15 | 0.351 | 0.049 | 4,916 | 1,493 |
|  |  | PHQ-2 | 1.26E-21 | 0.259 | 0.038 | 5,936 | 2,378 |
|  | Depression Cases | PHQ-8 | 4.30E-06 | 0.306 | 0.078 | 1,667 | 511 |
|  |  | PHQ-2 | 4.51E-08 | 0.217 | 0.063 | 2,070 | 789 |
| All of Us | All | PHQ-9 | 0.981 | 0.006 | 0.055 | 2,223 | 576 |
|  |  | PHQ-2 | 0.006 | 0.079 | 0.041 | 5,515 | 1,279 |
|  | Depression Cases | PHQ-9 | 0.129 | 0.018 | 0.044 | 4,421 | 951 |
|  |  | PHQ-2 | 0.922 | -0.02 | 0.058 | 1,960 | 491 |
| MGB | All | PHQ-9 | 1.55E-32 | 0.195 | 0.028 | 11,448 | 3,404 |
|  |  | PHQ-2 | 4.02E-90 | 0.281 | 0.022 | 16,696 | 5,446 |
|  | Depression Cases | PHQ-9 | 2.76E-26 | 0.173 | 0.029 | 10,782 | 3,165 |
|  |  | PHQ-2 | 9.70E-60 | 0.23 | 0.023 | 15,130 | 4,795 |
| Meta-analysis | All | PHQ-8 or 9 | 1.09E-18 | 0.196 | 0.022 | - | - |
|  |  | PHQ-2 | 3.91E-44 | 0.241 | 0.017 | - | - |
|  | Depression Cases | PHQ-8 or 9 | 8.51E-10 | 0.142 | 0.023 | - | - |
|  |  | PHQ-2 | 1.25E-22 | 0.198 | 0.020 | - | - |

PHQ-2 values also associated with antidepressant response. Worsening antidepressant response associated with a 0.47-point increase (beta = 0.26, SE = 0.04, p-value = 1.26 x 10^−21^) in PHQ-2 across all samples, and a 0.41-point increase among depression cases (beta = 0.22, SE = 0.06, p-value = 4.51 x 10^−8^) (Table 2, Supplementary Figure 2).

### Replication in the All of Us Research Program and MGB

We applied our antidepressant response algorithm to 92,614 individuals and 3,205,667 antidepressant records in the All of Us Research Program, resulting in 54,235 individuals and 1,991,779 response outcomes. The demographics of the All of Us sample were similar to the VUMC sample, with high proportions of female (68.7%), white (65.9%), and SSRI users (55.5%) (Table 1).

We extracted PHQ-9 and PHQ-2 results from All of Us and MGB and associated the change in PHQ over time with antidepressant response using longitudinal models. In All of Us, the PHQ-9 models included 576 individuals and 2,223 observations and the PHQ-2 models included 1,279 individuals and 5,515 observations. PHQ-9 was not associated with antidepressant response over time across sites (p-value = 0.981, beta = 0.01, SE = 0.06), however, PHQ-2 did associate with response (p-value = 0.006, beta = 0.08, SE = 0.04) (Figure 2, Table 2, Supplementary Figure 2). While the PHQ-9 longitudinal models did not align with the VUMC results, there were modest differences in mean PHQ-9 between response groups (responders: 12.6, intermediate – antidepressants only: 12.7, intermediate with antipsychotics: 15.0, non-responders: 13.8).

In MGB, the PHQ-9 models across all individuals contained 3,404 individuals and 11,448 observations, while the PHQ-2 models contained 5,446 individuals on 16,696 observations. Within MGB, PHQ-9 and PHQ-2 exhibited significant associations with antidepressant response over time for all individuals (PHQ-9: p-value = 1.55 x 10^−32^, beta = 0.20, SE=0.03; PHQ-2: p-value = 4.02 x 10^−90^, beta = 0.28, SE=0.02), consistent with results from VUMC. The effect estimates correspond to an increase of 1.3 points in PHQ-9 and 0.53 points in PHQ-2 with each level of worsening antidepressant response. Within depression cases, both PHQ-9 (p-value = 2.76 x 10^−26^, beta = 0.17, SE=0.03) and PHQ-2 models (p-value = 9.70 x 10^−60^, beta = 0.23, SE = 0.02) replicated associations seen in VUMC, corresponding to increases of 1.11 points in PHQ-9 and 0.44 points in PHQ-2. (Figure 2A, Table 2).

The meta-analysis across the three sites indicated worsening antidepressant response associated with increased PHQ-8 or PHQ-9 scores across all samples (p-value = 1.09 x 10^−18^, beta = 0.20, SE = 0.02) and within depression cases (p-value = 8.51 x 10^−10^, beta = 0.14, SE = 0.02). Worsening antidepressant response also associated with increased PHQ-2 scores in the meta-analysis of all individuals (p-value = 3.91 x 10^−44^, beta = 0.24, SE = 0.02) and depression cases (p-value = 1.25 x 10^−22^, beta = 0.20, SE = 0.02) (Figure 2B, Table 2).

### GWAS of response to first antidepressant trial

The GWAS of response to first antidepressant trial consisted of 21,644 responders, 2,437 intermediate – antidepressant only, 2,045 intermediate with antipsychotics, and 4,026 non-responders. No associations exceeded the statistical threshold for genome-wide significance (Supplementary Figure 3). The SNP-based heritability of the GWAS of antidepressant response was estimated as 3.84% (SE = 0.007).

The GWAS of antidepressant response was significantly genetically correlated with depression (rg = 0.23, SE = 0.06, p-value = 1.87 x 10^−4^), bipolar (rg = 0.15, SE = 0.05, p-value = 9.92 x 10^−4^), schizophrenia (rg = 0.19, SE = 0.05, p-value = 1.28 x 10^−4^), and cross-disorders (rg = 0.18, SE = 0.06, p-value = 1.10 x 10^−3^) (Supplementary Table 7). The positive correlation estimates indicate worse antidepressant response is positively correlated with common genetic liability to psychiatric traits.

### Associations with psychiatric polygenic scores

Within VUMC and MGB, we tested for associations between response to first antidepressant trial and polygenic scores for depression, anxiety, bipolar, schizophrenia, and cross-disorders. Among individuals of European ancestry, worse response to first antidepressant trial significantly associated with increased genetic liability for all tested psychiatric PGS (anxiety: p-value = 3.44 x 10^−4^, OR = 1.04; bipolar: p-value = 8.15 x 10^−7^, OR = 1.06; cross-disorders: p-value = 9.32 x 10^−10^, OR = 1.08; depression: p-value = 5.09 x 10^−18^, OR = 1.11; schizophrenia: p-value = 7.50 x 10^−7^, OR = 1.06).

After controlling for depression diagnosis, all PGS remained associated, except anxiety. Additionally, the odds ratios were slightly attenuated after controlling for depression diagnosis (bipolar: p-value = 1.99 x 10^−3^, OR = 1.04; cross-disorders: p-value = 1.03 x 10^−3^, OR = 1.05; depression: p-value = 2.84 x 10^−8^, OR = 1.08; schizophrenia: p-value = 5.93 x 10^−4^, OR = 1.05) (Figure 4, Supplementary Table 8).

There were no significant associations among individuals of African ancestry in VUMC (Supplementary Table 9, Supplementary Figure 4).

**Figure 3.**
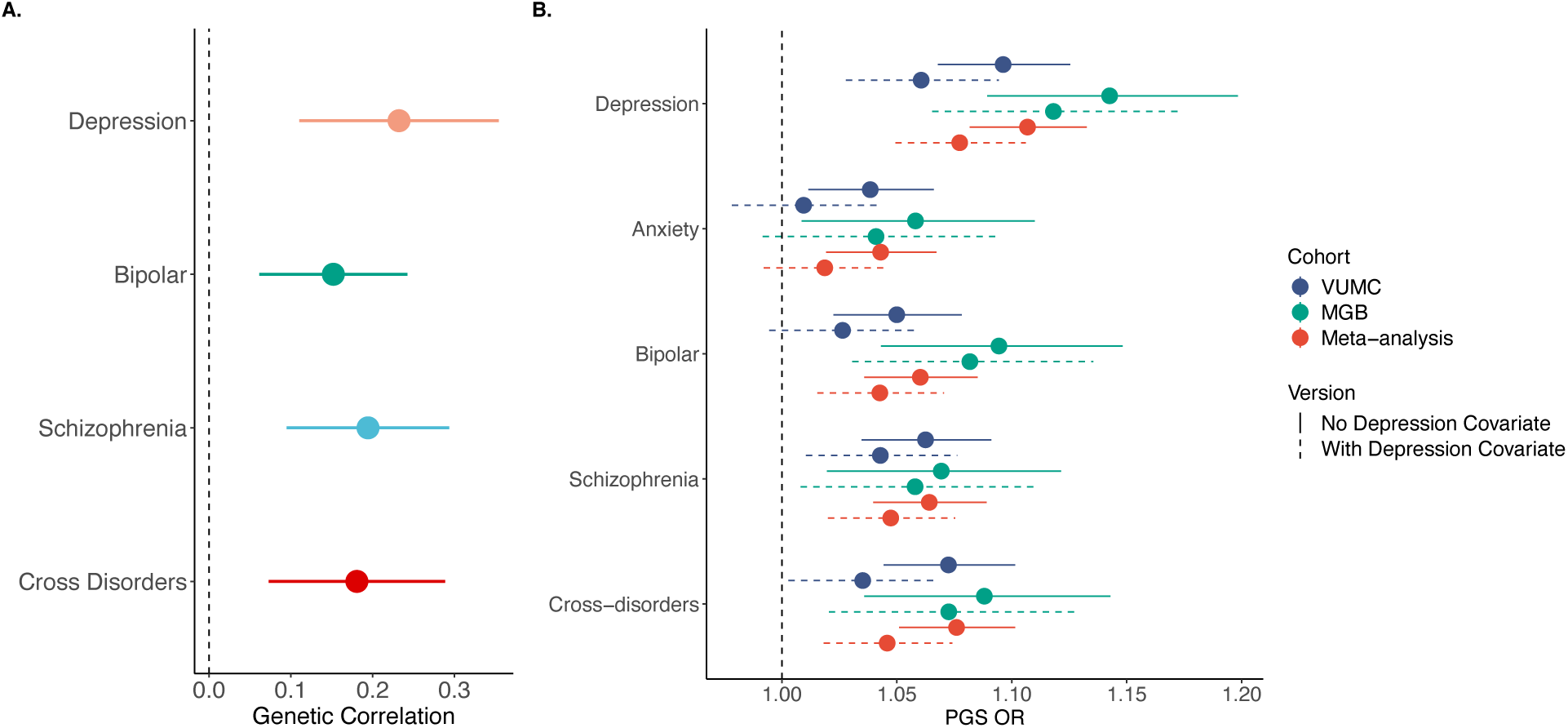
**A**. Genetic correlation between GWAS of response to first antidepressant trial in BioVU and psychiatric traits. **B**. Results of ordinal regressions between psychiatric polygenic scores and response to first antidepressant trial across cohorts and meta-analyzed in samples of European genetic ancestry. Dashed lines represent models controlled for depression diagnosis. All models were controlled for age at response, sex, and top 10 principal components of ancestry.

## Discussion

In this study, we presented an antidepressant algorithm using medications extracted from EHRs. Antidepressant response is poorly understood and identifying clinically actionable factors can help provide quicker relief from depressive symptoms for patients. EHRs provide a wealth of existing information on large numbers of antidepressant users to probe for clinically relevant factors associated with response at low cost.

We inferred antidepressant response based on medication switching or augmentation/combination therapy across three healthcare systems. When restricting to response to first antidepressant trial, our algorithm identified 77.5% of individuals as responders, greater than the previously reported 35% (ref). Because we do not have access to an individual’s lifetime medical history, it is possible that an appreciable proportion of individuals entered the healthcare systems already prescribed an antidepressant to which they responded favorably, potentially overrepresenting “responders” in our sample.

While our algorithm is easily transferrable across EHR systems, it does have several notable limitations. Our algorithm does not distinguish between switching due to non-response versus switching due to adverse effects of antidepressants. Pairing our algorithm with natural language processing algorithms to extract adverse drugs events from clinical notes could help distinguish between non-response and side effects. We also do not account for drug dosage in our antidepressant algorithm due to high levels of missingness in dosage data in EHRs. Medications prescribed or mentioned in EHRs does not guarantee a patient filled the prescription or takes the medication as prescribed.

Using two versions of the patient health questionnaire in VUMC, we confirmed our algorithm captured changes in depressive symptoms over time, with worse response from our algorithm associating with higher depressive symptom scores during treatment. The effect estimates correspond to a greater than 1-point increase in PHQ-8 and PHQ-9 scores with each increase in worsening antidepressant response class. Notably, the antidepressant-PHQ pairs used in the model are approximated by time between recorded dates, introducing some uncertainty between the matched PHQ score and inferred antidepressant response. In the future, mood symptom tracking recorded between visits and at visits for patients on antidepressants would help to rapidly and more accurately match depressive symptoms to response.

We replicated our algorithm in two different EHR datasets, the All of Us Research Program and MGB Healthcare System, to further validate the relationship between antidepressant response outcomes and PHQ. The association between antidepressant response and PHQ scores robustly replicated within MGB for both PHQ-9 and PHQ-2 scores, indicating our antidepressant response algorithm can be transferred between healthcare systems and PHQ versions. Within All of Us, the associations replicated with PHQ-2 scores, but not with PHQ-9 scores. However, there were group mean differences between the response groups, with responders and intermediate – antidepressants only clustering together and intermediate with antipsychotics and non-responders showing similar PHQ scores. The absence of replication in the All of Us data may stem from differences in biobank recruitment between All of Us and the academic medical centers of VUMC and MGB. Additionally, variability in the use of PHQ and prescribing guidelines across medical institutions within the All of Us dataset could contribute to the inconsistency, unlike the more uniform conditions in the single-institution analyses conducted at VUMC and MGB.

Using BioVU samples, we conducted a GWAS of antidepressant response using response to first antidepressant trial. While the GWAS is still underpowered to detect significant associations, we found evidence for a complex genetic architecture. Genetic correlation analysis between antidepressant response and psychiatric traits and identified significant genetic correlations with all tested psychiatric traits, indicating shared genetic architecture. Importantly, while the genetic predisposition to psychiatric disorders was also associated with response, the genetic correlation between the two was not equal to one, indicating there are unique genetic features underlying antidepressant response.

Polygenic scores analysis replicated previous findings that individuals with higher genetic liability to depression and schizophrenia have worse response to their first antidepressant trial compared to individuals of lower genetic liability(38–40). We identified additional associations between worse antidepressant response and increased liability to bipolar and cross-disorders.

In conclusion, we describe a portable antidepressant response algorithm that uses only EHR medication data and infers response variables over the course of treatment. Our response algorithm associated with changes in depressive symptoms over time and genetic analysis indicated shared genetic architecture between response and psychiatric traits. EHRs are a powerful resource for increasing power of genetic studies and our algorithm provides a new way to explore response at scale.

## Data Availability

Data collected from electronic health records are not available for public sharing.

## Acknowledgements

The All of Us Research Program is supported by the National Institutes of Health, Office of the Director: Regional Medical Centers: 1 OT2 OD026549; 1 OT2 OD026554; 1 OT2 OD026557; 1 OT2 OD026556; 1 OT2 OD026550; 1 OT2 OD 026552; 1 OT2 OD026553; 1 OT2 OD026548; 1 OT2 OD026551; 1 OT2 OD026555; IAA #: AOD 16037; Federally Qualified Health Centers: HHSN 263201600085U; Data and Research Center: 5 U2C OD023196; Biobank: 1 U24 OD023121; The Participant Center: U24 OD023176; Participant Technology Systems Center: 1 U24 OD023163; Communications and Engagement: 3 OT2 OD023205; 3 OT2 OD023206; and Community Partners: 1 OT2 OD025277; 3 OT2 OD025315; 1 OT2 OD025337; 1 OT2 OD025276. In addition, the All of Us Research Program would not be possible without the partnership of its participants. JWS and LKD were supported in part by NIMH R01 MH118233.

## Disclosures

JWS is a member of the Scientific Advisory Board of Sensorium Therapeutics (with options), and has received grant support from Biogen, Inc. He is PI of a collaborative study of the genetics of depression and bipolar disorder sponsored by 23andMe for which 23andMe provides analysis time as in-kind support but no payments.

## Notes

### Funding Statement

JMS was funded by 1F31MH124306-01A1.

### Author Declarations

The Vanderbilt University Medical Center Institutional Review Board approved this project (IRB#190418).

