## Supplemental Information for "Cross-EHR validation of antidepressant response algorithm and links with genetics of psychiatric traits"

Description of Cohorts.

*VUMC Electronic Health Records and BioVU*

Vanderbilt University Medical Center (VUMC) is a tertiary care center that provides inpatient and outpatient care in Nashville, TN. The VUMC EHR was established in 1990 and includes data on billing codes from the International Classification of Diseases, 9$^{th}$ and 10$^{th}$ editions (ICD-9 and ICD-10), Current Procedural Terminology (CPT) codes, medications, laboratory values, reports, and clinical documentation. The de-identified mirror of the EHR, numbers more than 3.2 million patient records.

In 2007, VUMC launched a biobank, BioVU, which links a patient's DNA sample to their EHR. The BioVU consent form is provided to patients in the outpatient clinic environments at VUMC. The form states policies on data sharing and privacy, and should a signature be obtained, makes any blood leftover from clinical care eligible for BioVU banking. The VUMC Institutional Review Board oversees BioVU and approved this project (IRB#190418). This protocol was deemed non-human subjects research because all information is de-identified.

*All of Us Research Program*

The All of Us program is an NIH funded study aimed at collecting genetic and EHR information on 1 million individuals in the United States. Within All of Us data, we extracted antidepressants and antipsychotics using the pre-build ATC concept terms (OMOP concept ID for antidepressants = 21604686 and antipsychotics = 21604490 & 21604557), mapped medication names to generic names, and applied the algorithm as described in VUMC. The algorithm in All of Us resulted in 1,991,779 response outcomes on 54,235 individuals. We defined depression status using the depression SNOMED code in All of Us (OMOP concept ID = 440383).

*Mass General Brigham Biobank*

The MGB Healthcare System is a tertiary care center in Boston, Massachusetts. The MGB EHR system contains information on in-patient and out-patient encounters, medications, and laboratory data. The data structure of MGB is more closely aligned to VUMC's EHR data, allowing us to extract antidepressants, antipsychotics, and depression cases as described in VUMC. Applying the antidepressant response algorithm in MGB resulting in response outcomes for 285,036 individuals.

Supplementary Table 1. Brand and generic names of antidepressants extracted from electronic health records in Vanderbilt University Medical Center.

| Class | Generic Name | Brand Name |
|---|---|---|
| SSRI | citalopram | Celexa |
| | escitalopram | Lexapro |
| | fluoxetine | Prozac |
| | | Sarafem |
| | | Rapiflux |
| | | Selfemra |
| | fluvoxamine | Luvox |
| | paroxetine | Brisdelle |
| | | Paxil |
| | | Pexeva |
| | sertraline | Zoloft |
| SNRI | desvenlafaxine | Khedezla |
| | | Pristiq |
| | duloxetine | Cymbalta |
| | | Drizalma |
| | | Sprinkle |
| | | Irenka |
| | levomilnacipran | Fetzima |
| | milnacipran | Savella |
| | venlafaxine | Effexor |
| TCA | amitriptyline | Elavil |
| | | Endep |
| | | Vanatrip |
| | amoxapine | Asendin |
| | clomipramine | Anafranil |
| | desipramine | Norpramin |
| | doxepin | Silenor |
| | imipramine | Tofranil |
| | nortriptyline | Pamelor |
| | | Aventyl |
| | protriptyline | Vivactil |
| Atypical | bupropion | Aplenzin |
| | | Forfivo |
| | | Wellbutrin |
| | | Zyban |
| | | Budeprion |
| | mirtazapine | Remeron |
| | nefazodone | Serzone |
| | vilazodone | Viibryd |
| | vortioxetine | Brintellix |
| | | Trintellix |
| MAOI | phenelzine | Nardil |
| | tranylcypromine | Parnate |

Supplementary Table 2. Brand and generic names of antipsychotics extracted from electronic health records in Vanderbilt University Medical Center.

| Generic name | Brand name |
|---|---|
| aripiprazole | Abilify |
| asenapine | Saphris |
| asenapine | Secuado |
| brexpiprazole | Rexulti |
| cariprazine | Vraylar |
| clozapine | Clozaril |
| clozapine | Fazaclo |
| clozapine | Versacloz |
| iloperidone | Fanapt |
| lumateperone | Caplyta |
| lurasidone | Latuda |
| olanzapine | Zyprexa |
| olanzapine | Lybalvi |
| olanzapine | Symbyax |
| paliperidone | Invega |
| pimavanserin | Nuplazid |
| quetiapine | Seroquel |
| risperidone | Risperdal |
| risperidone | Perseris |
| sulpiride | Dolmatil |
| ziprasidone | Geodon |

Supplementary Table 4. Mean and standard deviation of patient health questionnaire scores stratified by PHQ version and depression status.

| Cohort | Version | Group | Mean | SD |
|---|---|---|---|---|
| VUMC | PHQ-8 | All | 6.49 | 5.38 |
| | | Depression | 7.21 | 5.57 |
| | PHQ-2 | All | 1.77 | 1.80 |
| | | Depression | 1.94 | 1.88 |
| All of Us | PHQ-9 | All | 13.01 | 6.41 |
| | | Depression | 13.33 | 6.29 |
| | PHQ-2 | All | 1.98 | 1.98 |
| | | Depression | 2.16 | 2.00 |
| MGB | PHQ-9 | All | 9.54 | 6.53 |
| | | Depression | 9.78 | 6.54 |
| | PHQ-2 | All | 2.12 | 1.89 |
| | | Depression | 2.25 | 1.90 |

Supplementary Table 5. Sample sizes by antidepressant class and first trial response used for the genome-wide association scan in BioVU. Outcomes were ordered from lowest to highest as ‘responder’, ‘intermediate – antidepressants only’, ‘intermediate with antipsychotics’, and ‘non-responder’.

| **Response** | **SSRI** | **SNRI** | **TCA** | **Atypical** | **MAOI** | **Total** |
|---|---:|---:|---:|---:|---:|---:|
| Responder | 12,635 | 3,500 | 3,044 | 2,457 | 8 | 21,644 |
| Intermediate - antidepressants only | 1,090 | 421 | 526 | 400 | 0 | 2,437 |
| Intermediate with antipsychotics | 1,234 | 410 | 112 | 284 | 5 | 2,045 |
| Non-responder | 2,386 | 625 | 561 | 452 | 2 | 4,026 |
| Total | 17,345 | 4,956 | 4,243 | 3,593 | 15 | 30,152 |

Supplementary Table 6. Phecodes and component ICD codes used to define depression cases. Cases were required to have at least 2 component ICD codes for a given phecode on different days to quality as a depression case. Samples with only one code were excluded.

| **Depression (phecode 296.2)** | **Major Depression (phecode 296.22)** | **Adjustment Reaction (phecode 304)** | **Dysthymic Disorder (phecode 300.4)** |
|---|---|---|---|
| 296.2 | 296.2 | 309 | 300.4 |
| 296.2 | 296.2 | 309 | F34.1 |
| 296.21 | 296.22 | 309.1 | |
| 296.22 | 296.23 | 309.2 | |
| 296.23 | 296.24 | 309.22 | |
| 296.24 | 296.25 | 309.23 | |
| 296.25 | 296.26 | 309.24 | |
| 296.26 | 296.3 | 309.28 | |
| 296.3 | 296.3 | 309.29 | |
| 296.3 | 296.32 | 309.3 | |
| 296.31 | 296.33 | 309.4 | |
| 296.32 | 296.34 | 309.8 | |
| 296.33 | 296.35 | 309.82 | |
| 296.34 | 296.36 | 309.83 | |
| 296.35 | F20.4 | 309.89 | |
| 296.36 | F32.8 | 309.9 | |
| 311 | F33.0 | F43.2 | |
| F20.4 | F33.1 | F43.8 | |
| F32.8 | F33.2 | F43.9 | |
| F33.0 | F33.3 | F94.8 | |
| F33.1 | F33.4 | | |
| F33.2 | F33.8 | | |
| F33.3 | | | |
| F33.4 | | | |
| F33.8 | | | |

Supplementary Table 7. Results of genetic correlation analysis between response to first antidepressant trial and psychiatric disorders.

| **Trait** | **Rg** | **SE** | **p-value** |
|---|---:|---:|---:|
| Depression | 0.232 | 0.062 | 1.87E-04 |
| Bipolar | 0.152 | 0.046 | 9.91E-04 |
| Schizophrenia | 0.194 | 0.051 | 1.28E-04 |
| Cross Disorders | 0.181 | 0.055 | 1.01E-03 |

Supplementary Table 8. Results of ordinal regression between response to first antidepressant trial and psychiatric polygenic scores in VUMC, MGBB, and meta-analysis for samples of European genetic ancestry.

| Cohort | Version | Trait | p-value | Odds Ratio | Lower OR CI | Upper OR CI |
|---|---|---|---|---|---|---|
| VUMC | No Depression Covariate | Anxiety | 4.89E-03 | 1.04 | 1.01 | 1.07 |
| | | Bipolar | 3.20E-04 | 1.05 | 1.02 | 1.08 |
| | | Cross-disorders | 2.76E-07 | 1.07 | 1.04 | 1.10 |
| | | Depression | 6.82E-12 | 1.10 | 1.07 | 1.13 |
| | | Schizophrenia | 7.67E-06 | 1.06 | 1.03 | 1.09 |
| | With Depression Covariate | Anxiety | 0.557 | 1.01 | 0.98 | 1.04 |
| | | Bipolar | 0.106 | 1.03 | 0.99 | 1.06 |
| | | Cross-disorders | 0.033 | 1.04 | 1.00 | 1.07 |
| | | Depression | 2.37E-04 | 1.06 | 1.03 | 1.09 |
| | | Schizophrenia | 9.17E-03 | 1.04 | 1.01 | 1.08 |
| MGB | No Depression Covariate | Anxiety | 0.021 | 1.06 | 1.01 | 1.11 |
| | | Bipolar | 2.30E-04 | 1.09 | 1.04 | 1.15 |
| | | Cross-disorders | 7.70E-04 | 1.09 | 1.04 | 1.14 |
| | | Depression | 4.35E-08 | 1.14 | 1.09 | 1.20 |
| | | Schizophrenia | 0.006 | 1.07 | 1.02 | 1.12 |
| | With Depression Covariate | Anxiety | 0.105 | 1.04 | 0.99 | 1.09 |
| | | Bipolar | 0.001 | 1.08 | 1.03 | 1.14 |
| | | Cross-disorders | 0.006 | 1.07 | 1.02 | 1.13 |
| | | Depression | 5.86E-06 | 1.12 | 1.07 | 1.17 |
| | | Schizophrenia | 0.022 | 1.06 | 1.01 | 1.11 |
| Meta-analysis | No Depression Covariate | Anxiety | 3.44E-04 | 1.04 | 1.02 | 1.07 |
| | | Bipolar | 8.15E-07 | 1.06 | 1.04 | 1.09 |
| | | Cross-disorders | 9.32E-10 | 1.08 | 1.05 | 1.10 |
| | | Depression | 5.09E-18 | 1.11 | 1.08 | 1.13 |
| | | Schizophrenia | 1.50E-07 | 1.06 | 1.04 | 1.09 |
| | With Depression Covariate | Anxiety | 0.170 | 1.02 | 0.99 | 1.05 |
| | | Bipolar | 0.002 | 1.04 | 1.02 | 1.07 |
| | | Cross-disorders | 0.001 | 1.05 | 1.02 | 1.07 |
| | | Depression | 2.84E-08 | 1.08 | 1.05 | 1.11 |
| | | Schizophrenia | 5.93E-04 | 1.05 | 1.02 | 1.08 |

Supplementary Table 9. Results of ordinal regression between response to first antidepressant trial and psychiatric polygenic scores in VUMC for samples of African genetic ancestry.

| Ancestry | Version | Trait | pvalue | OR | Lower.CI | Upper.CI |
|---|---|---|---|---|---|---|
| AFR | No Depression Covariate | Anxiety | 0.776 | 1.01 | 0.94 | 1.09 |
| | | Bipolar | 0.721 | 0.98 | 0.90 | 1.07 |
| | | Cross-disorders | 0.652 | 0.98 | 0.88 | 1.08 |
| | | Depression | 0.875 | 0.99 | 0.92 | 1.08 |
| | | Schizophrenia | 0.941 | 1.00 | 0.91 | 1.09 |
| | With Depression Covariate | Anxiety | 0.64 | 1.02 | 0.93 | 1.12 |
| | | Bipolar | 0.73 | 0.98 | 0.89 | 1.09 |
| | | Cross-disorders | 0.34 | 0.94 | 0.84 | 1.06 |
| | | Depression | 0.95 | 1.00 | 0.91 | 1.10 |
| | | Schizophrenia | 0.69 | 0.98 | 0.88 | 1.08 |

Supplementary Figure 1. Histograms of number of timepoints per individual in PHQ-antidepressant models.

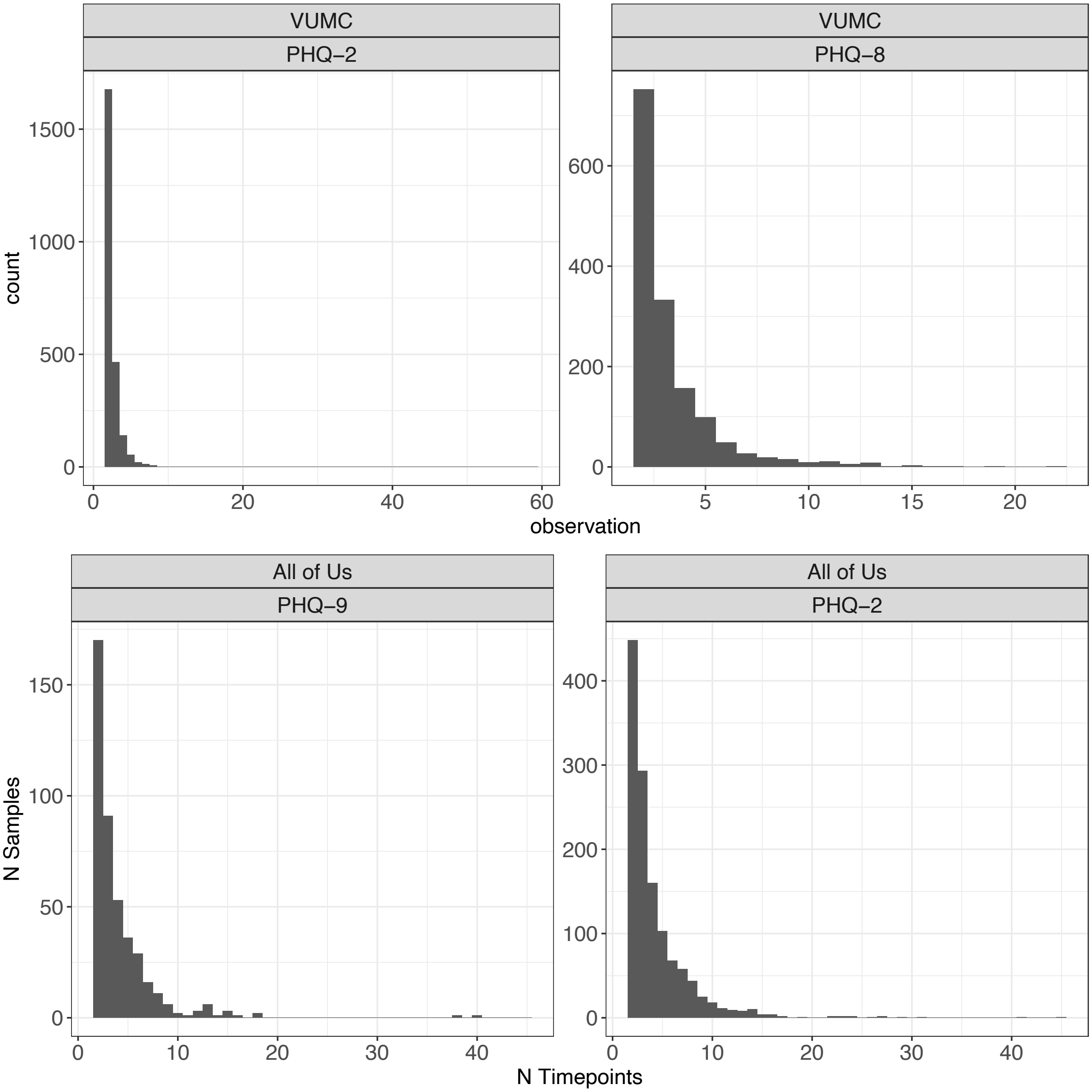

Supplementary Figure 2. Boxplot of PHQ-2 scores by antidepressant response in VUMC MGB, All of Us across A) all samples and B) depression cases.

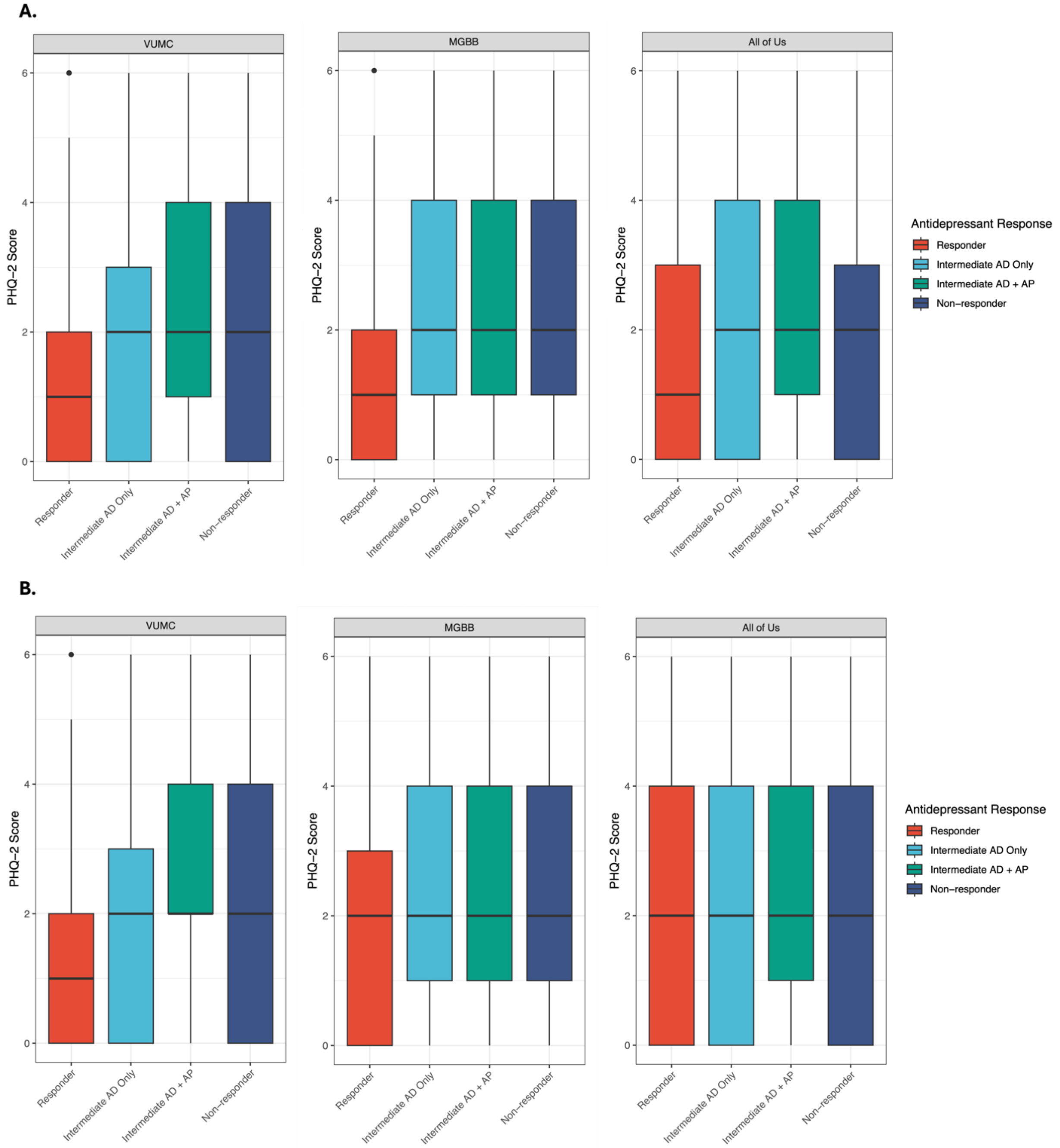

Supplementary Figure 3. A) QQ plot and B) Manhattan plot of p-values of the ordinal GWAS of response to first antidepressant response. The blue line on the Manhattan plot represents p-value = 0.05 and the red line indicates genome-wide significante (5 x $10^{-8}$).

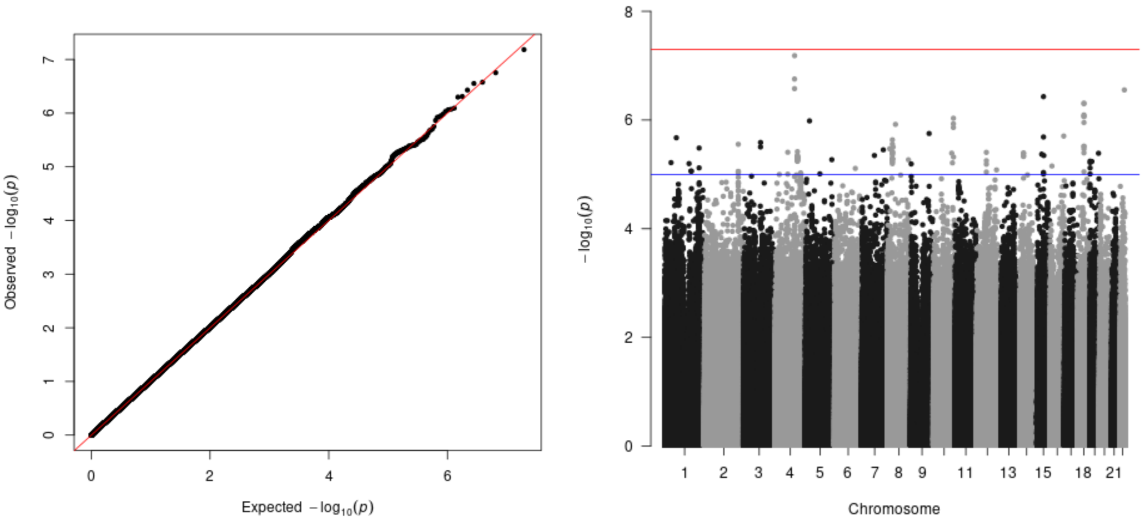

Supplementary Figure 4. Results of ordinal regression between response to first antidepressant trial and psychiatric polygenic scores in VUMC for samples of African genetic ancestry.

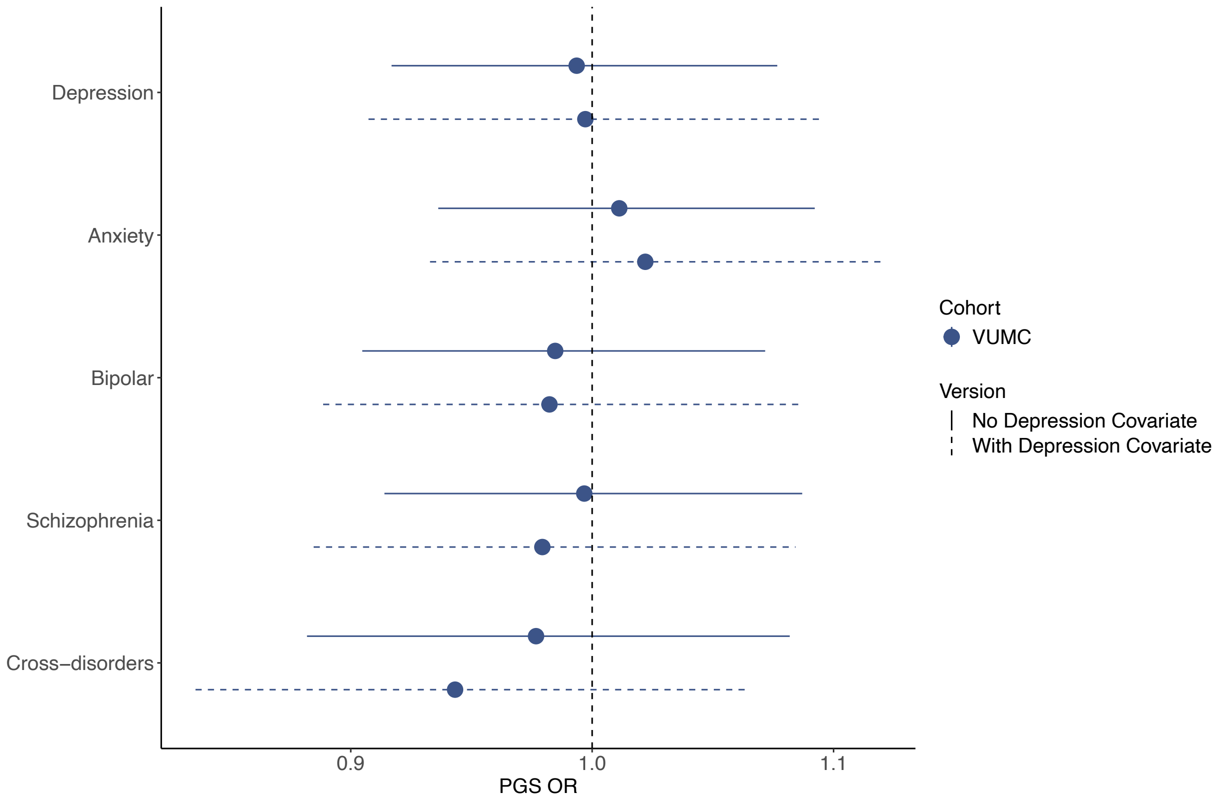
